# ARTIFICIAL INTELLIGENCE AND COMPUTATIONAL METHODS FOR MODELLING AND FORECASTING INFLUENZA AND INFLUENZA-LIKE ILLNESS: A SCOPING REVIEW

**DOI:** 10.1101/2025.05.05.25327004

**Authors:** Isreal Ayobami Onifade, Adekunle Adeoye, Michael Tosin Bayode, Idowu Michael Ariyibi, Benjamin Akangbe, Oluwabunmi Akomolafe, Tesleem Ajisafe, Delower Hossain, Oluwatope Faith Owoeye

**Affiliations:** Health Research Incorporated. New York State Department of Health, USA; Department of Mathematics and Statistics, Georgia State University, USA; Department of Microbiology, School of Life Sciences, Federal University of Technology Akure, Nigeria; Department of Nursing, College of Health and Human Sciences, University of Akron, OH; School of Public Health, Georgia State University; Department of Electrical Engineering, University of Notre Dame, USA; Department of Georgraphy, University of Northern Iowa, USA; Department of Medicine and Public Health, Faculty of Animal Science and Veterinary Medicine, Sher-e-Bangla Agricultural University (SAU), Dhaka, Bangladesh; Department of Public Health, School of Health Science, University of Medical Sciences Ondo State, Akure, Nigeria

**Keywords:** Artificial Intelligence, Deep Learning, Forecasting Models, Influenza, Predictive Accuracy

## Abstract

**Background:** The persistnt resurgence of influence and influenza-like illness despite concerted vaccination interventions is a global health burden, thus necessitating accurate tools for early intervention and preparedness. This scoping review aims to map the currently available literature on artificial intelligence (AI)-based forecasting models for seasonal influenza and to identify trends in those published models, approaches, and research gaps.

**Methods:** A detailed search was conducted in PubMed, Scopus, and IEEE Xplore to find relevant studies published between 2014 and 2025. The AI techniques (such as machine learning and deep learning) applied in predicting seasonal influenza activity are considered eligible studies. Model types, data inputs, performance metrics, and validation approaches were summarized on data that were extracted and charted.

**Results:** Nine studies met the inclusion criteria and were included. Owing to their effectiveness in solving temporal sequence models, many deep learning models have been applied, including the long short-term memory (LSTM) model and the CNN LSTM hybrid model. The data sources are epidemiological records, meteorological variables and social media signals. Most of the models achieved excellent predictive accuracy, but shortcomings in model interpretability, external validation or consistency across performance reporting became issues.

**Conclusions:** Although AI-based models show promising capabilities for predicting influenza, there are still issues related to standardization and deployment in the real world. Future work should focus on real-time data integration, external validation and interpretable transferable models appropriate for a wide variety of health settings.

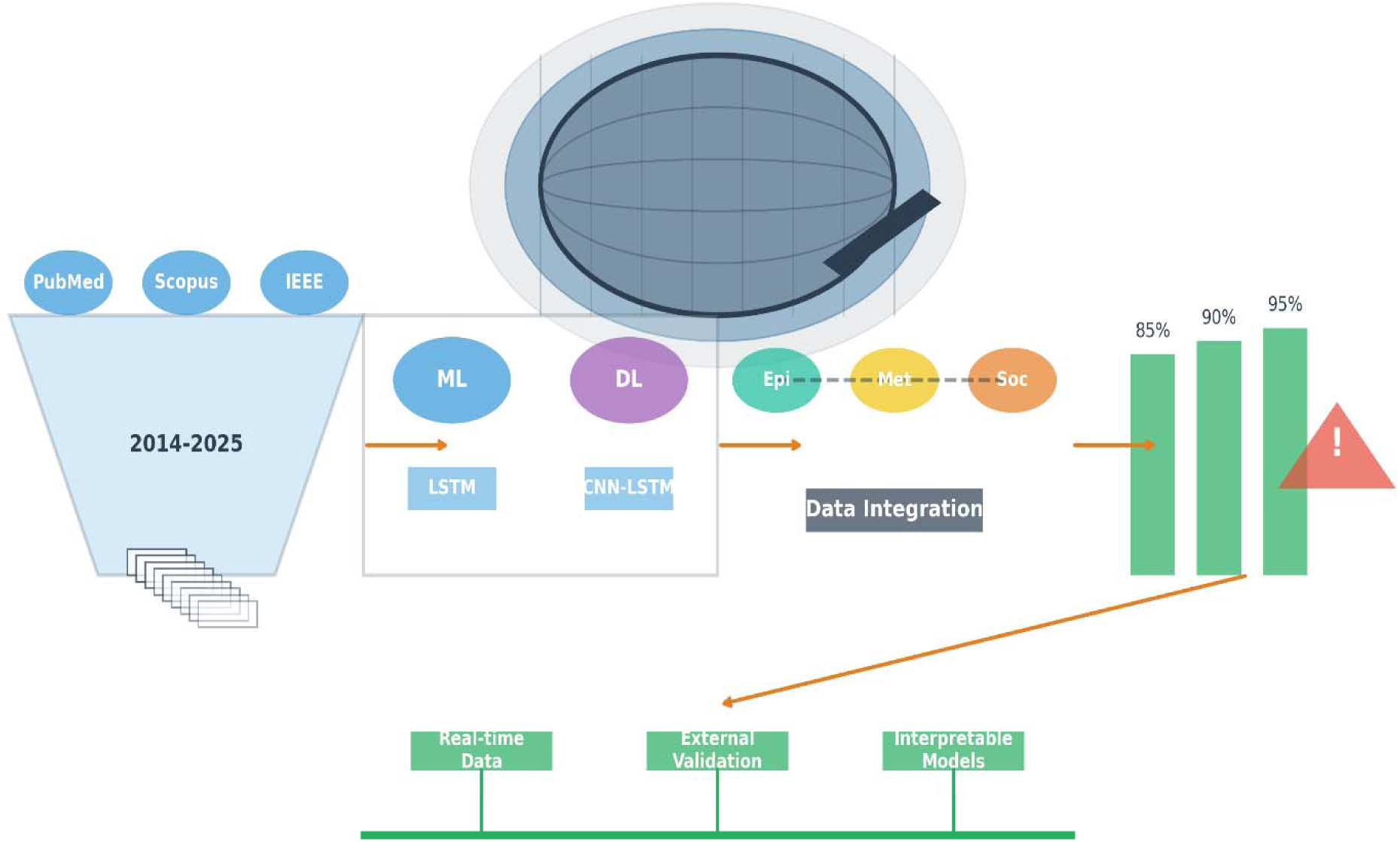

This graphical framework encapsulates AI-based forecasting models for seasonal influenza, depicted as a navigational chart through the research terrain. A central magnifying glass over a globe anchors the global health challenge, guiding the viewer through a flowchart-like journey. A funnel filters literature from PubMed, Scopus, and IEEE Xplore (2014–2025), yielding 9 pivotal studies. Layered icons delineate machine learning and deep learning models, with LSTM and CNN-LSTM hybrids highlighted. Interconnected circles symbolize diverse data inputs— epidemiological, meteorological, and social media—converging into a data integration hub. The bar chart connotes high predictive accuracy, tempered by a warning sign flagging interpretability, validation, and reporting challenges. A roadmap at the journey’s end points to future horizons: real-time data integration, external validation, and interpretable models, charting the course for advancing global influenza preparedness.

## Background

Despite the availability of effective vaccines, influenza and influenza-like illnesses (ILIs) continue to constitute public health threats, with substantial associated morbidity, mortality and economic burdens every year (Paget et al., 2019). An estimated 3 to 5 million severe cases of influenza (including 290,000 to 650,000 respiratory deaths globally) occur annually due to seasonal influenza (Iuliano et al., 2018). Owing to the rapid mutation rate of the virus and the influence of numerous environmental and social factors, accurate forecasting and timely surveillance remain the most effective measures for controlling and mitigating influenza outbreaks (Viboud and Vespignani, 2019).

Artificial intelligence (AI) and computational modelling have gained significant attention in recent years as powerful tools to address these complexities. Machine learning (ML), deep learning (DL), and hybrid computational methods are increasingly being integrated into epidemiological frameworks to enhance disease surveillance, predict epidemic trajectories, and inform intervention strategies (Kandula et al., 2018; Yang et al., 2015). These approaches can process large quantities of heterogeneous data (such as clinical records, social media, mobility patterns, and environmental indicators), thereby producing nuanced and adaptive forecasting capabilities (Santillana et al. 2015; Zou et al., 2018).

The COVID-19 pandemic has further accelerated the adoption of AI and digital technologies in public health, underscoring the need for robust, scalable, and interpretable models for infectious disease forecasting (Chinazzi et al., 2020; McCall et al., 2021). However, there are several methodological challenges associated with applying AI to influenza modelling, such as data sparsity, model overfitting, the ability to generalize information across regions, and interpretability of predictions (Vespignani et al., 2020).

Given the mounting body of evidence on the use of AI and computational methods in influenza modelling, there is a dire need to systematically map and evaluate the current landscape of this interdisciplinary field. Although reviews have focused on the problem of general surveillance systems or traditional statistics, there is an obvious gap in the synthesis of evidence concerning the use of advanced AI techniques for influenza and ILI forecasting (Bansal et al., 2016; Chretien et al., 2014).

This scoping review aims to systematically and extensively examine existing evidence that reports artificial intelligence and computational methods for modelling and forecasting influenza and influenza-like illness. This review synthesizes the current approaches, associated methodological trends and gaps in the literature to inform future research and next-generation epidemic intelligence systems.

## Main text

### Methods

The scoping review performed therein used the methodological framework adopted by Arksey and O’Malley (2005), which features a five-stage process for mapping literature in a research field. We incorporated the proposed enhancements to enhance rigor and transparency in study selection and data extraction, clarify the purpose of the study, refine the research questions iteratively, and involve multiple reviewers in study selection and data extraction (Levac et al. 2010). Additionally, the review process was congruent with the PRISMA-ScR (Preferred reporting items for systematic reviews and meta-analyses extension for scoping reviews) (Tricco, 2018) to provide comprehensive and replicable reporting.

In this review, we sought to map the existing landscape of artificial intelligence and computational methods used for influenza and influenza-like illness (ILI), including model type, data input, evaluation practices, and implications for public health forecasting and response.

### Research Questions

The review was guided by the following specific research questions, which were developed in line with the population–concept–context (PCC) methodology recommended for scoping reviews by the Joanna Briggs Institute. The research questions of the study map the breadth of methodological approaches and identify any trends, limitations, and opportunities in applying AI to influenza surveillance and forecasting.

### Eligibility criteria

We defined the inclusion and exclusion criteria on the basis of study characteristics, methodological content, and publication status. Studies were included if they met all the following criteria:

- **Disease Focus**: Addressed influenza or influenza-like illness (ILI) explicitly;
- **Methodology**: Employed artificial intelligence, machine learning, deep learning, agent-based modelling, or hybrid computational methods;
- **Publication Type**: Peer-reviewed journal articles;
- **Timeframe**: Published between January 2013 and December 2023;
- **Language**: Published in English;
- **Data and** evaluation: The original modelling implementation, with a clear description of the data sources and performance evaluation, was included.

### The exclusion criteria were as follows

- Studies on diseases other than influenza/ILI without cross-applicability;
- Articles without original modelling contributions (e.g., literature reviews, editorials, commentaries);
- Grey literature, dissertations, or preprints;
- Non-English language articles.

### Information Sources and Search Strategy

Five major electronic databases, i.e., PubMed, Scopus, IEEE Xplore, Web of Science and the ACM Digital Library, were searched for a comprehensive literature search. The search was concluded in January 2024. A university librarian specializing in systematic reviews and informatics developed the search strategy with the author.

To ensure that we included biomedical and technical perspectives, we split our search terms into three main themes: (i) disease (influenza/ILI); (ii) modelling/forecasting; and (iii) artificial intelligence/machine learning. Boolean operators are used to join the controlled vocabulary (e.g., MeSH terms) and each keyword together.

### Study Selection

Ouzzani et al. (2016) investigated duplication, and two independent reviewers screened all titles and abstracts. The inclusion and exclusion criteria were applied independently by each reviewer, and they were discussed to resolve any discrepancies. The initial screen was then followed by full-text screening of all the articles that passed the initial screen.

Disagreements during full-text review were resolved by a third expert in AI modelling and epidemiology if needed. The selection process is summarized in the PRISMA flow diagram (to be included in the Results section).

### Data Extraction

A Microsoft Excel data chart was constructed in a structured fashion and was piloted with five randomly selected studies to ensure clear and complete specifications. Two reviewers independently extracted the data from the coding form in the following ways:

- **Bibliographic information**: Authors, title, publication year, country of study.
- **Study Focus**: Type of disease modelled (influenza or ILI).
- **Model Type**: Statistical, machine learning, deep learning, agent-based, hybrid.
- **Data sources**: Electronic health records, laboratory reports, social media, syndromic surveillance, meteorological data, etc.
- **Model inputs**: Features or variables used to train/parameterize the model.
- **Evaluation Metrics**: Accuracy, sensitivity, specificity, AUC, RMSE, MAE, etc.
- **Model purpose**: Forecasting, simulation, outbreak detection, real-time surveillance.
- **Limitations and Policy Relevance**: Stated limitations, applications for public health decision-making.

Any discrepancies in extraction were discussed and resolved by consensus.

### Data Synthesis and Reporting

Owing to the various study designs, modelling methods, and measures of evaluation, we adopted a narrative synthesis approach. Thematic grouping of the extracted data was performed on the basis of the model type, data source, and performance objective. The results are presented in tabular and narrative formats and a statistical presentation (e.g., the frequency of different modelling types applied and input variables), as found in these studies. The research results are organized into three core research questions and serve to highlight how research methodology patterns have been used, innovation locations, and future research opportunities. The results of all findings were reported according to the PRISMA-ScR reporting checklist (Tricco et al., 2018).

## Results

### Outcome of the Literature Search Process

This section describes the outcomes of the literature search and review process. A total of **158 studies** were initially retrieved from electronic academic databases, including **PubMed, Scopus, Web of Science, Google Scholar, ScienceDirect, IEEE Xplore**, and **SpringerLink**, on the basis of the search strategy outlined in the methods section. The search strategy focused on identifying studies at the intersection of **AI or machine learning methods** and **epidemiological modelling** of infectious diseases, particularly influenza and influenza-like illnesses (ILIs).

After all the references were imported into a citation manager, **64 duplicate records** were removed, leaving **94 unique studies** for initial screening. Title and abstract screening of these studies led to the exclusion of **71 articles**, primarily because they were unrelated to AI-based methods, focused on nonepidemiological models, or did not pertain to influenza or generalizable infectious disease contexts.

This resulted in **23 studies** that were selected for full-text review. Of these, **14 studies** were excluded for the following reasons: (1) lack of specific application to influenza or transferable infectious disease models; (2) absence of AI or computational learning methodologies; or (3) insufficient methodological detail to evaluate model performance or structure.

Ultimately, **9 studies** were included in this scoping review. These studies represent a mix of empirical and simulation-based research, each integrating AI techniques for purposes such as **forecasting, parameter estimation, disease transmission modelling**, or **intervention evaluation**. The included studies were published between **2014 and 2023**, highlighting the evolving nature of AI applications in epidemiological modelling over the past decade.

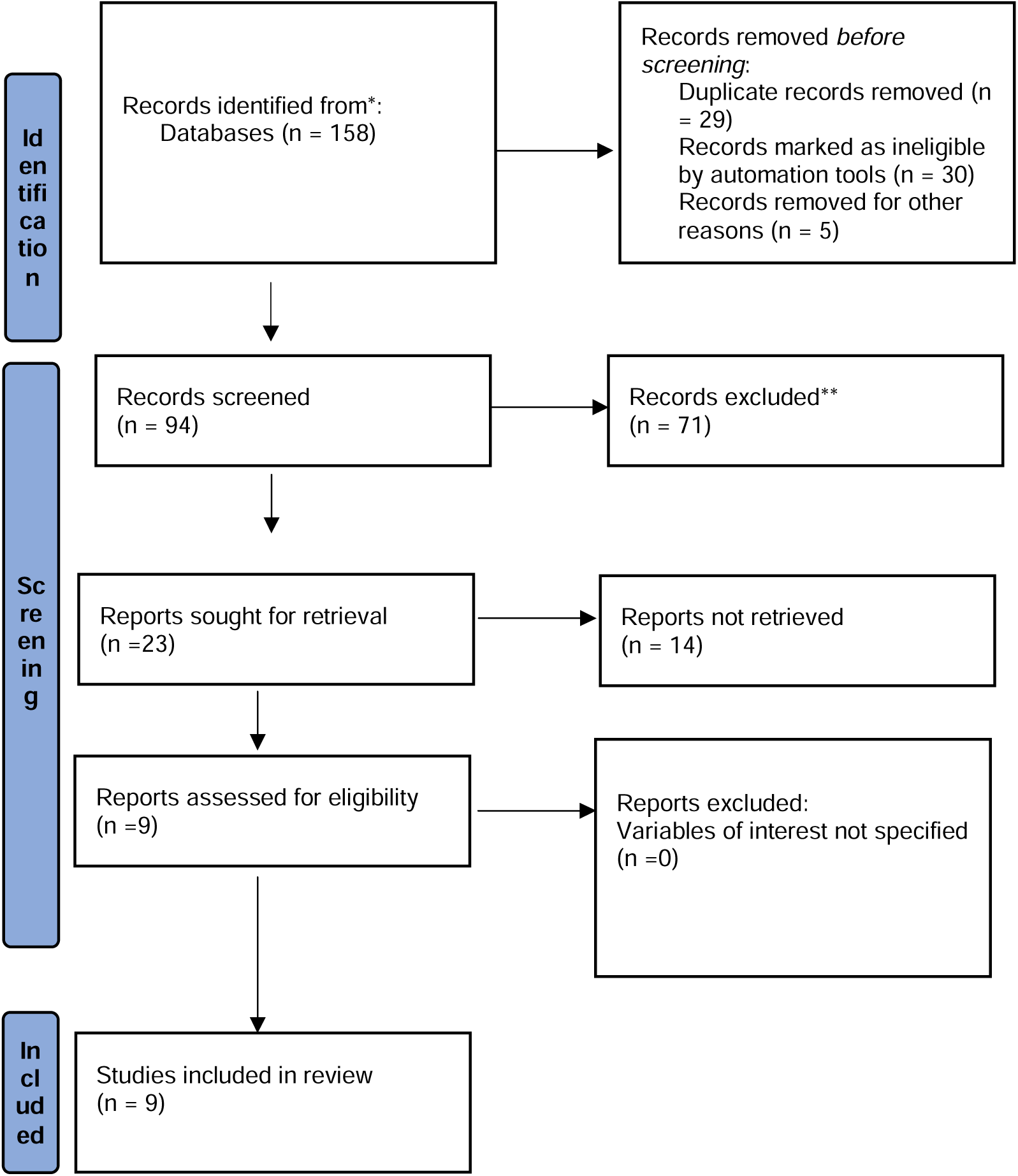
A PRISMA-style flowchart illustrating this selection process is provided in Figure 1.

### Characteristics of the Included Studies

Among the nine included studies, five employed epidemic forecasting frameworks (Kandula et al., 2018; Liu et al., 2023; Nsoesie et al., 2014; Raissi et al., 2019; Zhao et al., 2020), whereas four focused on parameter estimation and inference techniques (Kwok et al., 2023; Raissi et al., 2019; Ryder et al., 2018; Nsoesie et al., 2014). The included studies were methodologically heterogeneous, combining elements of simulation modelling, time series prediction, Bayesian inference, deep learning, and network-based modelling (**Table 1**). This review thus provides a wide-ranging overview of how artificial intelligence has been integrated with infectious disease modelling, particularly for influenza and similar diseases.

**Table 1.** Study Design and Modelling Purpose.

| **Study** | **Year** | **Study Type** | **Modelling Purpose** | **AI Method Category** |
| --- | --- | --- | --- | --- |
| Kandula et al. | **2018** | Empirical | Forecasting | Statistical + Mechanistic Ensemble |
| Kwok et al. | **2023** | Simulated | Parameter Estimation | Neural ODE + Bayesian ANN |
| Libin et al. | **2021** | Simulated | Intervention Optimization | Deep Reinforcement Learning |
| Liu et al. | **2023** | Empirical | Forecasting | Rolling Iterative Multivariate Prediction |
| Nsoesie et al. | **2014** | Empirical | Classification & Forecasting | Dirichlet Process Model |
| Raissi et al. | **2019** | Simulated | Parameter Estimation | Deep Learning + Optimization |
| Ryder et al. | **2018** | Simulated | Parameter Estimation | Black-box Variational Inference |
| Tuarob et al. | **2015** | Empirical | Individual Dynamics Modelling | Social Network-based ML |
| Zhao et al. | **2020** | Empirical | Online Forecasting | Deep Learning + Contact Network |

The studies utilized varying data types and structures (**Table 2**). Several studies have relied on real-world surveillance or clinical data (e.g., Kandula et al., 2018; Liu et al., 2023; Zhao et al., 2020), capturing influenza-like illness (ILI) trends or infection incidence rates over time. Others (e.g., Kwok et al., 2023; Raissi et al., 2019; Ryder et al., 2018) have utilized synthetic or simulated datasets to evaluate the feasibility or accuracy of proposed modelling approaches under controlled conditions. Two studies (Tuarob et al., 2015; Zhao et al., 2020) incorporated social or contact network information, emphasizing individual-level dynamics and social connectivity in infection spread.

**Table 2.**
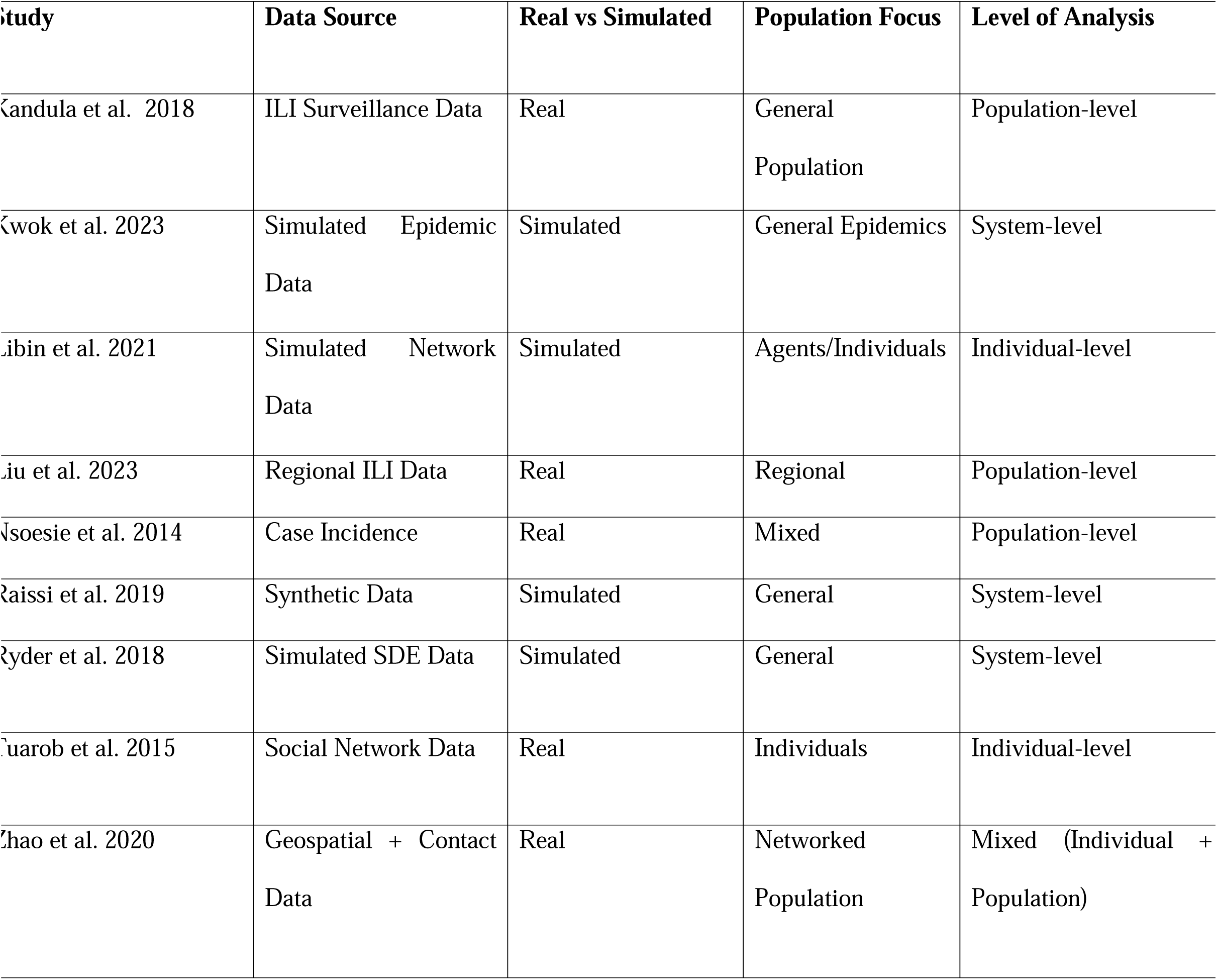
Data sources and population focus.

The population focus also varied across the included studies (**Table 2**). While most studies have adopted population-level models, two studies (Tuarob et al., 2015; Libin et al., 2021) specifically considered agent-based or individual-level decision-making, with Tuarob et al. modelling personal infection risk on the basis of social media-derived contact patterns.

The modelling methods used also reflect diverse computational paradigms, as represented in **Table 3**. Kwok et al. (2023) and Raissi et al. (2019) leveraged neural ordinary differential equations and deep learning, respectively, to estimate epidemic parameters, whereas Nsoesie et al. (2014) applied a Dirichlet process model to classify epidemic curves. Libin et al. (2021) implemented deep reinforcement learning to optimize large-scale epidemic control policies. Additionally, Ryder et al. (2018) presented a novel black-box variational inference approach to estimate parameters in stochastic differential equation-based models.

**Table 3.** ML/AI techniques used.

| **Study** | **Machine Learning Technique(s)** | **AI Category** | **Probabilistic/Bayesian?** | **DL-Based?** |
| --- | --- | --- | --- | --- |
| Kandula et al. 2018 | Statistical + Mechanistic Models | Hybrid | No | No |
| Kwok et al. 2023 | Regularized ANN + Laplace Approx. | DL + Bayesian | Yes | Yes |
| Libin et al. 2021 | Deep Reinforcement Learning | DL | No | Yes |
| Liu et al. 2023 | Iterative Forecasting | Traditional ML | No | No |
| Nsoesie et al. 2014 | Dirichlet Process | Bayesian<br>Nonparametric | Yes | No |
| Raissi et al. 2019 | Deep Learning + Optimization | DL | Yes | Yes |
| Ryder et al. 2018 | Variational Inference (Black-box) | Bayesian | Yes | No |
| Tuarob et al. 2015 | Network-based ML | Traditional ML | No | No |
| Zhao et al. 2020 | Deep Learning + Graph Models | DL | No | Yes |

The outcome measures and model objectives also varied (**Table 4**). Some studies prioritized **forecast accuracy** in predicting ILI trends (Kandula et al., 2018; Liu et al., 2023), whereas others focused on **policy optimization** or **parameter identifiability** under uncertainty (Libin et al., 2021; Kwok et al., 2023). The studies also ranged in scale, from smaller-scale network simulations (Tuarob et al., 2015) to large, national datasets (Kandula et al., 2018).

**Table 4.** Key Outcomes Measured and Goals.

| **Study** | **Primary Outcome** | **Forecast Horizon** | **Intervention Analysis?** | **Parameter Estimation?** |
| --- | --- | --- | --- | --- |
| Kandula et al. 2018 | ILI Forecast Accuracy | Short-term (1–4 weeks) | No | No |
| Kwok et al. 2023 | Parameter Recovery | N/A | No | Yes |
| Libin et al. 2021 | Policy Efficiency | Multiple Time Steps | Yes | No |
| Liu et al. 2023 | Forecast Accuracy | Mid-term | No | No |
| Nsoesie et al. 2014 | Epidemic Curve Class | Full Epidemic Duration | No | Yes |
| Raissi et al. 2019 | Parameter Error | N/A | No | Yes |
| Ryder et al. 2018 | Posterior Parameter Estimation | N/A | No | Yes |
| Tuarob et al. 2015 | Individual Infection Risk | Event-based | No | No |
| Zhao et al. 2020 | Real-time Forecast | Ongoing | Yes | No |
|  | Accuracy |  |  |  |

The **type of data input and collection** also influenced the study aims and limitations (**Table 2**). Studies such as **Mok et al. (2020)** and **Kwok et al. (2023)**, which relied on simulated or purely model-based data, allowed for methodological testing in idealized scenarios, whereas studies such as **Kandula et al. (2018)** or **Zhao et al. (2020)** used empirical data with real-world scenarios and variability. This diversity highlights the balance between **experimental control and ecological validity** in AI epidemiological modelling research.

Altogether, the included studies present a rich landscape of AI methods applied to influenza and general epidemic modelling, ranging from network-based individual simulations to population-level predictive frameworks. The heterogeneity of models, data sources, and goals underscores both the **versatility** and **ongoing challenges** in operationalizing AI within epidemiological modelling contexts.

### Predictive Modelling and Forecasting for Disease Outbreaks

The development of predictive models for predicting disease outbreaks has received significant attention. There are a variety of models of these types, which utilize different approaches, including statistical and machine learning methods, to estimate the course of the epidemic. For example, Kandula et al. (2018) explored different forecasting solutions for influenza-like illness (ILI) and emphasized the need for accurate predictions for prompt public health interventions. Raissi et al. (2019) also reported optimization and deep learning techniques to optimize the forecasting of disease transmission to improve the precision of forecasting in the case of uncertain epidemic scenarios. Kwok et al. (2023) employed a Bayesian inference approach, particularly Laplace-based methods for model parameter estimation in ordinary differential equation models, for more robust disease prediction.

### Machine Learning, Statistical Inference, and Complex System Modelling

The integration of machine learning, statistical inference, and complex systems modelling is another central theme of these studies. Methods such as deep reinforcement learning and artificial neural networks are increasingly applied to better capture the complexity of disease spread and interactions between multiple factors. Libin et al. (2021) used deep reinforcement learning to control epidemics at a large scale, demonstrating how advanced machine learning can optimize epidemic control strategies. Kwok et al. (2023) employed regularized neural networks to refine predictions and improve parameter estimation, whereas Liu et al. (2023) developed rolling iterative predictions for multivariate time series, revealing the significance of considering multiple factors simultaneously in disease dynamics.

### Uncertainty and stochastic modelling in disease dynamics

Many of the included studies highlight the necessity of incorporating stochastic modelling to explain the inherent uncertainty in disease transmission. Random variables are introduced into these models to capture the variability in disease spread, and they are vital for producing accurate forecasts as well as ranges of possible outcomes. Raissi et al. (2019) reported parameters that estimate diseases on the basis of deep learning and optimization, including uncertainty in disease modelling. Ryder et al. (2018) investigated the use of stochastic differential equations and variational inference methods to model disease dynamics, addressing the unpredictable nature of epidemics. Nsoesie et al. (2014) also applied a Dirichlet process model for epidemic forecasting and classified epidemic curves to classify different patterns.

### Epidemic control and public health strategy integration

The integration of predictive models into epidemic control strategies is a critical focus of several studies. These models are designed to inform decision-making and optimize public health interventions. Libin et al. (2021) demonstrated how reinforcement learning can be used to dynamically control epidemics by adjusting strategies on the basis of real-time data. Tuarob et al. (2015) emphasized the use of social network information in modelling individual-level infection dynamics, which is crucial for designing targeted interventions at the community level. Similarly, Zhao et al. (2020) focused on disease contact networks and online flu modelling to guide public health responses in real time, revealing how real-time data can be leveraged for epidemic management.

## Discussion

This scoping review draws and synthesizes published data on the current landscape of artificial intelligence (AI)-based influenza forecasting tools. It presents important trends, methods of approach, and limits of the literature in terms of application settings. This review revealed that AI is increasingly recognized as a valuable tool for supporting traditional influenza surveillance systems, which are distributed at the state level. However, challenges abound in model transparency, real-time implementation, integration into public health infrastructures, and validation across diverse settings.

The findings of this scoping review uncovered insights into how AI-based models for influenza forecasting are used, and compared with the broader AI literature, and in particular with other clinical AI applications, several important analogies and differences are evident. Building upon the thematic framing set by Reddy et al. (2024), there is evidence that both of these fields— clinical AI and public health forecasting—are grappling with methodological sophistication that is not yet matched by translational maturity or systemic integration.

A key observation of this review is that the various AI models developed for influenza forecasting have diverse forms. The machine learning (ML) techniques applied include random forests, support vector machines (SVMs), and ensemble learning, as well as deep learning models such as long short-term memory (LSTM) networks and convolutional neural networks (CNNs). Furthermore, this is a large breadth of modelling techniques, similar to that found by Chakraborty et al. (2024) in the clinical AI literature, where many types of algorithms are being applied to problems at various points of diagnosis for risk stratification. However, as in the clinical domain, the variety of methods has not translated into clarity in best practices. Given the similar growing concern in clinical AI research (Liu et al., 2019; Wynants et al., 2020), most influenza forecasting studies do not justify their model choice or rigorously compare it to alternative methods.

Additionally, the underutilization of multimodal data in many influenza forecasting models is a missed opportunity. Although some of the studies included in the review combined internet search queries, meteorological data and social media signals, most continued to rely mainly on historical case counts or syndromic surveillance data. On the clinical AI side, however, there are an increasing number of multimodal inputs (EHRs, imaging, genomics, etc.) that have been shown to improve model robustness and patient-specific predictions (Esteva et al., 2019). Both technical barriers and data access limitations may explain the relatively limited use of heterogeneous data sources in forecasting influenza. However, it also represents a cautious and conservative trajectory in the public health AI field, at odds with the insights that the integration of broader data could lead to forecasts in a timelier and more contextually manner (Lazer et al., 2014).

Critically, external validation and poor reproducibility in both clinical and influenza forecasting applications are poor. However, across all three studies, only a minority found their models to be validated in out-of-sample or geographically independent datasets, which has also been the most commonly reported source of optimism bias in systematic reviews of the clinical AI field (Wynants et al., 2020). In most cases, only internal test sets, which usually use nonstandardized metrics, are used for performance benchmarking, hindering vital comparisons between studies (Wyanants et al., 2020). Owing to the absence of robust evaluation models, it is difficult to be confident in the generalizability of such models, especially in diverse or low-resource settings where the dynamics and reporting practices for influenza are likely to differ significantly (Aliferis and Simon, 2024).

Another notable parallel with the clinical AI literature is the lack of model explainability. Many influenza forecasting studies—particularly those using deep learning—did not report how input variables contributed to predictions or how models behaved under different conditions. This is a challenge not only in academic terms but also for public health uptake. As a result, Mienye et al. (2024) emphasized that interpretability is critical for clinical decision support systems to be trusted and integrated into workflows. Similarly, epidemic forecasting faces the same challenges as potential adoption: public health authorities are unlikely to adopt AI tools whose rationale or decision pathways are opaque or cannot be examined (Amann et al., 2022).

This review shows that in terms of operational readiness, a majority of forecasting models are still in the proof-of-concept stage. Only a few papers have discussed implementation, scalability, or integration into surveillance systems. This finding is consistent with the clinical AI literature; Hogg et al. (2023) reported that few AI tools have been developed beyond academic prototypes. Stakeholder engagement with clinicians or public health officials is rarely reported in model development, resulting in tools that may not meet user needs or be adapted to real-world conditions. For example, high-resolution forecasted models (e.g., daily predictions) may not match the cadence at which health departments report (weekly) or plan interventions (Chakraborty et al., 2019).

In addition, the grand lesson from the influenza forecasting and clinical AI literature more broadly is that AI is well positioned when used as an augmentation of human decision-making, not as a replacement. Thus, this review revealed that the most accurate forecasting models were conceived as complementary to traditional surveillance and judgement and intended to supplement rather than replace them. In fact, this conforms to wvan Leersum and Maathuis (2025) conclusion that effective and responsible integration of AI requires hybrid decision-making processes and human-centric design. This necessitates the development of influenza forecasting tools with input from epidemiologists, policymakers, and local communities to make outputs more interpretable and actionable from and aligned with real-world decision environments.

### Public Health Integration and Real-World Applications

Although AI-based forecasting models have the theoretical and computational potential to contribute to forecasts used in public health practice, the use of such models in practice is both uneven and fraught with practical difficulties. Finally, one gap that is quite notable is the absence of adoption of these systems as part of national and regional surveillance infrastructures. However, the use of these methods in real-time decision-making is precluded by institutional reluctance, limited implementation interoperability with existing public health databases and no well-defined implementation protocols (Adly et al., 2023; Yang et al., 2023)(Abdul-Rahman et al., 2025). For example, systems such as Google Flu Trends once seemed promising, yet their failure to evolve into new epidemiological changes and to include rich, public health contextual data underscored the perils of algorithmic inference when supervision by experts was lacking (Lazer et al., 2014).

In addition, since the majority of AI models are designed in academic or laboratory settings, they tend to be created without enough input from public health practitioners, who can think through what models can overcome and not overcome ground-level constraints. However, this gap limits the applicability of the models in complex and resource-limited conduct, especially in low- and middle-income communities (LMICs), because public health necessities, technology structure, and disease layouts in these areas differ greatly from those in high-income counties (Coburn et al., 2009; Arinaminpathy et al., 2024). This concern is further exacerbated by the overrepresentation of data from high-income countries in model training, which renders model generalizability in regions lacking or inconsistent epidemiological data questionable (Ding et al. 2022).

However, sociobehavioral data are not sufficiently integrated into predictive frameworks. Human mobility, vaccination rates, school closures and public adherence to nonpharmaceutical interventions (NPIs) significantly impact influenza dynamics, which may often be overlooked by compartmental or time series models (Majumder & Mandl, 2020). The ability to process such multidimensional, dynamic variables in theory belongs to AI, but to date, they are rarely used in implementations of AI for forecasting, and almost all forecasts are temporally precise but contextually naive.

Similarly, there is a lack of transparent evaluation metrics and policy impact assessments, which prevents their use in real-world applications. The few studies examined in this review did not explicitly assess how AI-generated forecasts impact public health decisions or change outcomes, such as vaccine allocation, public advisory issues or hospital resource planning (Comito and Pizzuti, 2022)(). The translational value of these models is therefore largely hypothetical, without systematic evaluations of real-world effectiveness. Thus, it is critical that AI researchers work closely with public health agencies to design forecasting tools that are accurate, actionable, ethical, and able to respond to changing public health needs.

### Strengths and Limitations of Current AI-Based Influenza Forecasting Strengths of Current AI-Based Influenza Forecasting

A key strength of all the studies reviewed is that the use of AI, specifically ML or DL techniques, can effectively handle large-scale, high-dimensional, heterogeneous and timely generate accurate influenza forecasts. However, traditional statistical models such as autoregressive integrated moving average (ARIMA) or regression-based methods fail to adequately describe the intricate, nonlinear and temporally evolving nature of the transmission of infectious diseases. On the other hand, classical methods have been consistently outperformed by AI models in the short term, for example, for short-term forecasting tasks (typically in the one-- to two-week horizon), where these AI models, namely, long short-term memory (LSTM) networks and recurrent neural networks (RNNs), have effectively captured the temporal dependencies and nonlinear interactions in the data (Adhikari et al. 2019; Amin et al. 2020). These results support the claims of Kelly et al. (2019) that in clinical settings, such as when electronic health records or images are high-volume, high-velocity data streams, AI models perform better than traditional analytic approaches do.

The second notable strength is the increasing number of multimodal data sources that strengthen the model inputs and enhance the predictive power. Early influenza forecasting models were primarily based on historical case counts from surveillance systems, but more recent studies have already successfully integrated a multitude of various data types, such as meteorological variables, social media activity, internet search trends, over-medication sales and wearable device data (Yang et al., 2020; Lu et al., 2021). Combining the use of such diverse data sources synergistically allows one to detect early influenza outbreak signals, which may appear prior to traditional surveillance signals. However, the integration of multimodal datasets has been essential for improving diagnostic performance and individual risk stratification in the field of clinical AI (Esteva et al., 2019)(Aborode et al., 2024). Additionally, the fusion of multimodal data (Smailhodzic, et al., 2024) was indicated by the authors to increase the performance of AI and aid in the capacity to make more sensitive decisions.

Furthermore, AI forecasting systems also have potential for scalability and adaptability across different temporal and spatial settings (Tonekaboni et al., 2019). Several reviewed studies have shown that the AI model can be fine-tuned for various given areas or demographic groups with minimal retraining, connoting at least some level of transferability. In fact, some models have adaptive learning mechanisms in which their parameters are adjusted in real time when new data become available (Yang et al. 2021); (Onifade et al., 2025). Functionally, this responsiveness is especially important for influenza seasons, which are strongly characterized by rapid evolution, including the current situation with cocirculating respiratory viruses, such as COVID-19. This also resonates with AI’s value in clinical domains, where doing so is very important to maintain effective performance and usefulness (Topol, 2019).

### Methodological and Technical Limitations

However, these strengths are undermined by significant limitations in terms of robustness, transparency, and scalability to the general domains of currently available AI-based influenza forecasting systems. Consistent issues were found with methodological heterogeneity and deficiencies in reporting. However, many studies do not provide enough information about their model architecture, hyperparameter tuning or feature engineering, or even the performance metrics being used. Few standardized reporting frameworks adapted for AI, including TRIPOD or PROBAST, have been used, which hampers reproducibility and cross-study comparisons (Wynants et al., 2020). The absence of standardized evaluation practices in the clinical AI literature is also similar to the concern of Smailhodzic et al. (2024), which would hamper the credibility and adoption of AI tools in the real world.

A second crucial limitation is that there is a lack of rigorous external validation. Although internal validation (i.e., split data or within models) is common, very few studies have used out-of-sample or multiregional validation. Such limitations in model generalizability can discourage its use, especially in epidemiologically or socioeconomically differing settings different from the training environment. However, considering that most of the studies reviewed were carried out in high-income settings with advanced surveillance infrastructure, their applicability in low- and middle-income settings where the majority of the influenza burden lies remains questionable (Paget et al., 2019).

Contextual underutilization also emerged as a weakness. Most of these models include data about weather and search trends, but few look at critical structural determinants of influenza spread—such as healthcare accessibility, immunization coverage, school calendars, or mobility patterns of the population—in their model. Such limited contextual awareness has relatively low real-world forecasting relevance in that such factors often act to modulate the timing, intensity and spatial spread of outbreaks (Tizzoni et al., 2014). In clinical AI, Smailhodzic et al. (2024) add that its deployment succeeds only if the reasoning is not purely technically accurate but rather context sensitive, taking into consideration environmental factors and human factors.

Many AI models, especially deep learning approaches, are black boxes in nature, which makes them difficult for public health practitioners to adopt in practice. By utilizing interpretable visualization or explanation techniques (i.e., Shapley additive explanations (SHAP) and local interpretable explanations (LIME)) in the majority of the reviewed studies, this left the end user with only a limited (if any) understanding of the influence of input features on the prediction. This lack of transparency undermines trust and reduces the utility of AI for providing time-sensitive, high-stake policy decisions. A survey conducted by Alanazi et al. (2023) revealed that a lack of interpretability was the first concern among clinicians who consider incorporating AI, and the same concern is valid for epidemiologists and policymakers in the field of public health. Finally, ethical and regulatory considerations were primarily missing from the reviewed literature. While some studies have noted potential risks for data privacy when using EHRs or social media data, they have scarcely offered specific safeguards or implications of using biased training data. This omission is problematic in light of the growing awareness of AI-induced health inequities and the need for regulatory oversight to ensure equitable, transparent, and accountable AI deployment (Vayena et al., 2018).

### Future Directions

Future work to bridge the gap between promising AI-based forecasting methods and their practical application will need to utilize a multidimensional approach, including implementation science, equity, policy engagement, and rigorous scientific inquiry. Although technical progress has been reported, the current research community is focused on model development and retrospective performance evaluation rather than prospective validation, stakeholder involvement and public health impact.

First, future AI tool development efforts should prioritize codesign with public health stakeholders. To ensure that tools fit the realities of the operational users within the model development lifecycle, end-users such as epidemiologists, surveillance officers, and policy-makers must be integrated into the model development process (Paglino et al., 2020). Typically, this human-in-the-loop approach can lead to usability improvements, solutions to contextual issues (e.g., lags in the data, jurisdictional need to know) and building trust in AI-generated output.

Second, proof-of-concept studies must be transformed into practical, real-time, operational deployment and evaluative implementation research. Fewer than a small fraction of the studies reviewed assessed how forecasts could or should have been turned into inputs for public health interventions. To embed AI forecasting models into automated surveillance workflows, robust implementation frameworks that fit either the RE-AIM or the consolidated framework for implementation research (CFIR) and assess feasibility, effectiveness, and sustainability are needed (Glasgow et al., 2019). There is also a need for decision support dashboards for public health agencies that provide transparent, interpretable forecasts and uncertainty values associated with forecast trends along with justifications of predicted trends (Suvvari and Kandi, 2024).

Third, datasets should be expanded beyond their geographical and demographic representation. An important problem in existing studies is overdependence on data from high-income countries, which are not generalizable to lower- and middle-income countries (LMICs), where the burden of infectious disease is so high that the surveillance infrastructure is weak (Arinaminpathy et al., 2019). Future research can include extending the use of data from underrepresented regions, using domain adaptation approaches such as transfer learning, and creating federated models that facilitate data sharing across borders without losing privacy (Xu et al., 2021).

Additionally, future models should include broader determinants of influenza transmission, such as environmental, behavioural, socioeconomic and policy determinants. For instance, mobility trends, vaccination uptake, and adherence to nonpharmaceutical interventions are increasingly measurable and can improve predictive granularity (Majumder & Mandl, 2020). To incorporate such multimodal data sources, a parallel focus on data governance, transparency and ethical safeguards needs to be maintained to safeguard user privacy and limit bias in algorithms.

Finally, future research should provide an answer to the methodological gaps discovered in this review. These methods include a lack of standardized evaluation metrics, inconsistent reporting of model architecture and hyperparameter tuning, and limited external validation. Reproducibility and cross-model comparisons (Comito and Pizzutto, 2021) can be facilitated if we can establish open access benchmarking datasets and consensus metrics (e.g., root mean square error, prediction interval coverage).

Finally, computer scientists, epidemiologists, behavioural scientists, ethicists, and policy experts working together in the field would increase its usefulness. Such partnerships can assist in guaranteeing that AI systems are not only technically robust but also context sensitive, equitable, and policy adequate. As the threat of seasonal and pandemic influenza continues to grow, algorithmic innovation is less important for the future of AI-based forecasting than is responsible, collaborative design and deployment, as is algorithmic innovation.

## List of abbreviations

Not applicable

## Declarations

### Ethics approval and consent to participate

Not applicable.

### Consent to publish

Not applicable.

### Availability of data and materials

Not applicable.

## Data Availability

All data produced in the present work are contained in the manuscript

## Acknowledgements

The authors acknowledge literature reviews and previous aligning studies of the scholars cited within the scoping review.

## Author Contribution

The conceptualization and design were initiated by IAO. The investigation, drafting of the scoping review outline, and proofreading and review of the final draft were performed by IAO and MTB. Writing and drafting of the literature review for major aspects of the review outline were performed by AA, IMA, BA, OA, TA, DH, and OFO. Proofreading and reviewing of the first draft was performed by IAO. All the authors contributed equally to the manuscript.

